# Metatranscriptomic analysis of wastewater sites reveals a high abundance of antimicrobial resistance genes from hospital wastewater

**DOI:** 10.1101/2024.02.03.24302270

**Authors:** Sebastian Musundi, Samuel Ng’ang’a, Esther Wangui Wanjiru, Kate Sagoe, Ernest Wandera, Bernard Kanoi, Jesse Gitaka

## Abstract

**Objective:** This study analyzed the metatranscriptome of wastewater samples from different sites with a focus of identifying antibiotic resistance genes and the bacterial community.

**Methods:** Twenty-four wastewater samples were collected from a hospital, university sewer, and the influent and effluent of a wastewater treatment plant (WWTP). The metatranscriptome was sequenced to identify antimicrobial resistance genes (ARGs), mobile genetic elements, and bacterial community composition.

**Results:** Metatranscriptome analysis revealed varying abundances of ARG transcripts across different sites, with 84, 27, 14, and 9 ARG transcripts identified in wastewater collected from the hospital, university, influent, and effluent of a WWTP, respectively. Notably, hospital wastewater contained clinically relevant beta-lactam ARGs, including *bla-NDM-1* and several *blaOXA* transcripts. Four core ARGs, against sulfonamides *sul1* and *sul2* and aminoglycosides *aph(6)-ld* and *aph(’3’)-lb* were also identified. The predominant bacterial community comprised of *Gammaproteobacteria,* with priority pathogens like *Neisseria gonorrhea* and *Helicobacter pylori* present in hospital wastewater and WWTP influent.

**Conclusion:** These findings provide insights into the wastewater resistome and how meta-transcriptomic data can be utilized for the surveillance of antibiotic resistance. Overall, our study highlights the utility of wastewater surveillance in understanding and addressing the dissemination of antibiotic resistance and emphasizes the crucial role of proper wastewater management in protecting public and environmental health.

## 1. Introduction

Antimicrobial resistance (AMR) remains one of the major global health challenges of the 21^st^ century and has been prioritized by the World Health Organization as one of the top 10 threats to humanity worldwide. In 2019, drug-resistant infections accounted for approximately 4.95 million deaths globally and disproportionately affected low- and middle-income countries in sub-Saharan Africa [1]. To tackle the threat posed by AMR effectively, continuous surveillance, which is critical for understanding trends in pathogen incidence and monitoring the effectiveness of various interventions, needs to be carried out [2]. Large networks sharing AMR data at the national, regional, and global levels, including the Global Antimicrobial Resistance Surveillance System (GLASS), Central Asian and European Surveillance of Antimicrobial Resistance (CAESAR), and European Antimicrobial Resistance Surveillance Network (EARS-Net), have been established [3]. However, these bodies primarily focus on individual-level sampling with selection bias toward antibiotic-resistant bacteria (ARB) in clinical settings and do not represent AMR within the population [4]. Furthermore, most laboratory settings within low- and middle-income countries (LMICs) lack the infrastructure for patient-level surveillance; therefore, data from these countries are underrepresented despite the region facing the greatest threat posed by AMR.

Wastewater surveillance (WWS) is a cost-effective approach for monitoring the presence of ARB and ARGs at the population level, as it involves the biomass of an entire population of a community [5]. The success of using WWS was demonstrated during the SARS-CoV-2 pandemic, when it was used to provide real-time information about the circulating strains of the virus [6]. Currently, the use of WWS is quickly gaining traction in AMR surveillance since it provides insights into current and previous antibiotic use, real-time information about circulating ARB and an early warning system for the emergence of multidrug-resistant pathogens in the community [7]. Furthermore, information obtained from WWS can be used to evaluate the efficacy of various methods of treating wastewater before release into the environment. For example, previous research has shown that sewage treatment does not eradicate ARG or antibiotics, which can enter the food chain using sewage sludge and water in agriculture [8,9]. WWS provides a suitable approach for detecting ARB and ARGs and can complement other existing surveillance methods, especially within LMICs.

Surveillance sites for assessing the presence of ARGs and ARB include areas where antibiotics are primarily used, such as in clinical or hospital settings and wastewater treatment plants where disinfection occurs. Wastewater treatment plants (WWTPs) are hot spots for AMR since antibiotics from different sources, including those originating from healthcare settings, agriculture, and industry, converge [10]. Failure to remove ARGs and ARB from the effluent of WWTPs results in their release and spread to the environment horizontally through mobile genetic elements [11]. Apart from WWTPs, hospital settings have the highest use of antibiotics due to their heavy use, and their effluent is expected to harbor high numbers of ARB and ARGs [12]. One common unexplored area for antibiotic use is within residential or institutional settings, especially in low- and middle-income countries where the prescription of antibiotics is not strictly regulated. Considering that antibiotic use is common due to self-medication and overall misuse, evaluating wastewater that originates not only from common hot spots such as WWTPs and hospital settings but also from residential or institutional settings may provide insights into the spread of ARGs.

Recent advances in high-throughput sequencing technologies have provided a better understanding of the wastewater resistome, allowing for the rapid identification of ARGs and microbial communities. One such approach is the analysis of the wastewater metatranscriptome, which provides information about the active bacterial community and ARGs by quantifying the messenger RNA of specific genes [13]. Therefore, through meta-transcriptomic analysis of wastewater samples, we can determine the abundance and expression patterns of specific ARGs within various settings and shed light on the measures needed to control ARGs from wastewater sources. In this study, we identified ARGs across four different sites, including a hospital, university wastewater and the influent and effluent of a WWTP. Our findings revealed many ARGs in hospital effluent and from universities and fewer ARGs in the influent and effluent of WWTPs. Furthermore, we identified clinically relevant ARGs from hospital wastewater and identified mobile genetic elements and virulence factors that play a role in the horizontal transfer of ARGs.

## 2. Materials and Methods

### 2.1 Sample collection

Twenty-four wastewater samples were collected from the hospital, university hostels, influent, and effluent of a WWTP using Moore swabs, created by cutting cotton gauze into pieces of approximately 90 cm long and 30 cm wide. These cut pieces were folded three times, firmly tied at the center with a nylon string, placed in the inflow stream of wastewater, and fastened with the nylon string by tying them to a hook. After 24 hours, Moore swabs were retrieved, placed in Ziploc bags, transported to the laboratory on ice in a cooler box at -4°C, and stored in a -80°C freezer until processing.

### 2.2 RNA extraction and sequencing

The Moore swabs were processed by thawing overnight at +4°C after adding 50 mL of 1X PBS. Once thawed, the swab was squeezed into a beaker, and the liquid was transferred into two 50 mL Falcon tubes. Fifteen milliliters of the squeezed-out raw wastewater was centrifuged at 10,000 × g for 10 minutes. Ten milliliters of clear supernatant was retrieved and subjected to total nucleic acid extraction using the MagMAX™ Wastewater Ultra Nucleic Acid Isolation Kit with Virus Enrichment (A52610; Thermo Fisher Scientific™, Massachusetts, U.S.), following the manufacturer’s instructions. The extracted nucleic acids were eluted in 50 µL of elution buffer and stored at -80°C until further processing. The RNA integrity and quality were assessed using an Invitrogen Qubit 4 Fluorometer (Thermo Fisher Scientific, Inc.). The RNA was subjected to Illumina MiSeq next-generation sequencing as described previously [14]. A 200 bp fragment library ligated with barcoded adapters was constructed using NEBNext Ultra RNA Library Prep Kit for Illumina v1.2 (New England Biolabs) according to the manufacturer’s instructions. The cDNA library was purified using Agencourt AMPure XP magnetic beads (Beckman Coulter). After assessing the quality and quantity of the purified cDNA library, nucleotide sequencing was performed on an Illumina MiSeq sequencer (Illumina) using a MiSeq Reagent Kit v3 (Illumina).

### 2.3 Reads processing

Paired-end reads generated from RNA sequencing were demultiplexed and retrieved from the MiSeq platform. The reads were processed using an in-house Nextflow pipeline (https://github.com/sebymusundi/metatranscriptomics). Briefly, the read quality was assessed using FastQC, adapter sequences were removed using FastP [15,16], and low-quality reads were removed using Trimmommatic (SLIDING WINDOW = 4:15, MINLEN = 30) [17]. The clean reads were merged using PEAR, mapped to the human reference genome Hg38 using Bowtie2, and unmapped reads were filtered out using Samtools [18]. SortmeRNA was subsequently used to remove ribosomal RNA (rRNA) from the remaining reads using default parameters against several SILVA rRNA databases: silva-euk-18s-id95.fasta, silva-bac-23s-id98.fasta, silva-bac-16s-id90.fasta, silva-arc-23s- id98.fasta, silva-arc-16s-id95.fasta, rfam-5s-database-id98.fasta, rfam-5.8s-database-id98.fasta, and silva-euk-28s-id98.fasta [19].

### 2.4 De novo assembly and Read Quantification

The non-ribosomal reads were coassembled de novo based on their collection site using the default settings in Trinity [20]. The transcript files were indexed and mapped to individual samples using Bowtie2. The generated SAM file was converted to a BAM and subsequently sorted and indexed. Samtools idxstats was used to determine the number of mapped and unmapped reads and the length of each transcript. Relative gene abundance was calculated using a similar approach to reads per kilobase per million reads (RPKM). Briefly, relative abundance was calculated as RNum_Gi=Num_Gi/∑i=1nNum_Gi, where Num_Gi represents the proportion of the number of mapped reads for each specific gene or transcript (Gi) relative to the total number of mapped reads across all genes or transcripts in the sample (∑i=1n(Num_Gi)) as previously described (Hu et al., 2013).

### 2.5 Identification of antibiotic resistance genes

The presence of ARGs was predicted by the Resfinder database [22]. A transcript was considered an ARG if the similarity and query coverage were greater than 80%. Transcripts associated with ARGs were retrieved, and a plot comparing the relative abundance of each class and type of ARG was generated in RNum_Gi.

### 2.6 Identification of mobile genetic elements and virulence genes

The transcript files were further examined for the presence of different mobile genetic elements. The presence of plasmids and insertion sequences was predicted using the mobile genetic element finder version 1.0.3 [23]. Virulence factors were predicted in Abricate using the latest updated version of the VFDB database, with a percentage similarity and coverage of 80% [24].

### 2.7 Taxonomy classification

The Kraken2 database was downloaded to classify transcripts based on their taxonomy [25]. Kraken2 was run on the transcripts using the standard database released in March 2023 (https://genome-idx.s3.amazonaws.com/kraken/k2_standard_16gb_20231009.tar.gz). The WHO list of priority pathogens was retrieved from the generated Kraken report.

## 3. Results

### 3.1 Identification of Antibiotic Resistance Genes

All major ARGs against the antibiotic class aminoglycosides, beta-lactams, macrolides, macrolides, streptogramins, nitroimidazoles, phenicols, rifampicin, sulfonamides, tetracyclines, trimethoprim, and quinolones were detected in the hospital wastewater (Figure 1). The university wastewater had ARGs against tetracycline, sulfonamides, macrolides, beta-lactams, and aminoglycosides. The influent and effluent from the WWTP contained fewer classes of ARGs than wastewater from the hospital and university. ARGs against aminoglycosides, macrolides, sulfonamides, and tetracyclines, and aminoglycosides, macrolides, and sulfonamides, were present in the influent and effluent of the WWTP respectively (Figure 1). ARGs against tetracycline and macrolide genes were the most abundant from the referral hospital effluent, while ARGs against sulfonamides were predominant in the university wastewater.

**Figure 1.**
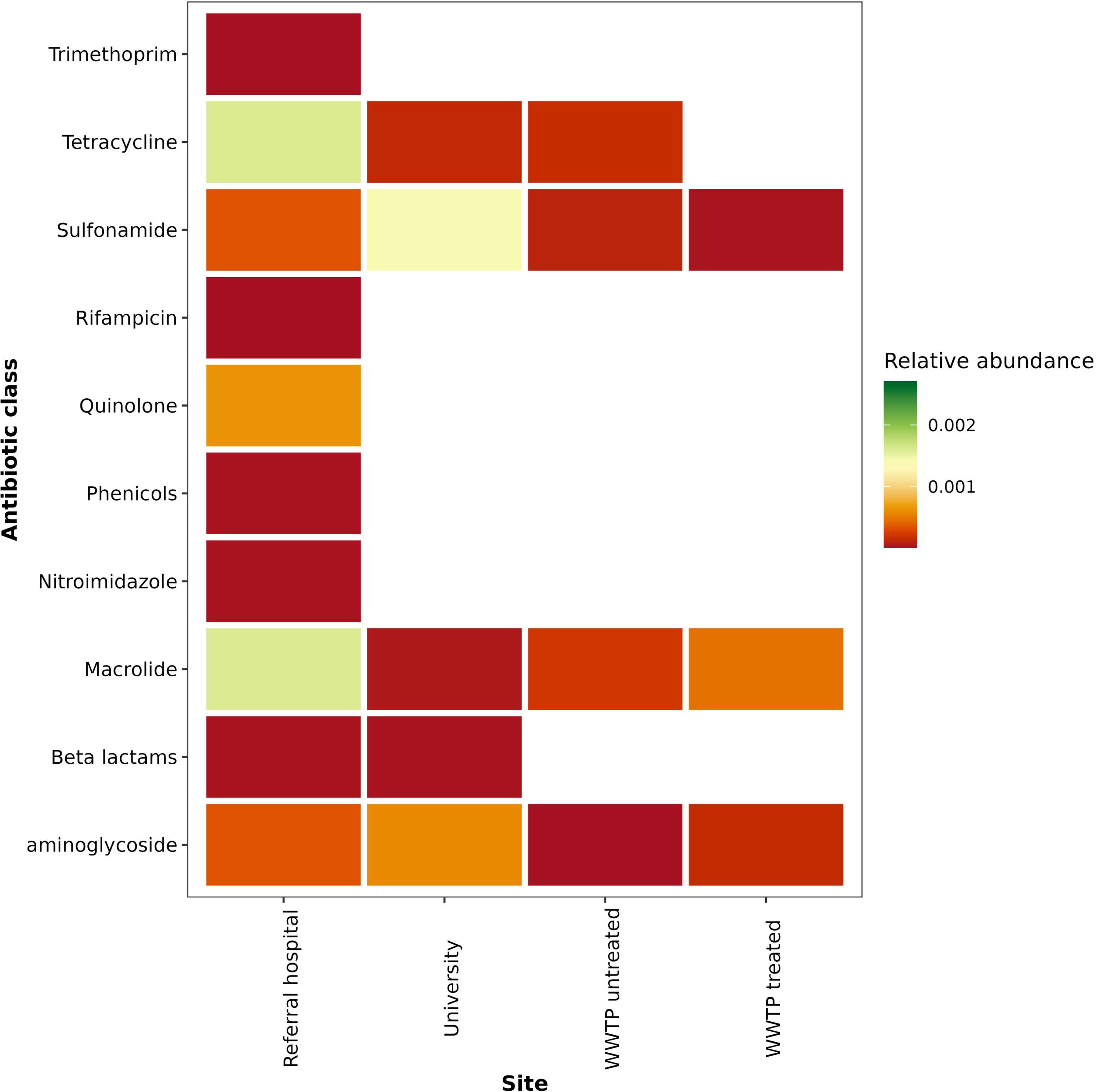
Antibiotic classes detected across the four different sites

We detected 84, 27, 14, and 9 ARG transcripts in the wastewater from the hospital, university, WWTP influent and effluent, respectively. A high number of transcript copies were identified for *sul2, sul1, msr(E), aph(3)-lb, aph(6)-ld, aac(6’)-lb-Hangzhou, mph(E), tet(Q), tet (W),* and *tet(39)*(Figure 2). The core antibiotic genes identified across all the sites included those against the sulfonamides *sul1 and sul2* and the aminoglycosides *aph(6)-ld* and *aph(’3’)-lb* (Figure 2). ARG genes that were present at 3/4 of the sites included *ant(2)-la, msr(E),* and *tet39*. ARGs at 2/4 of the sites were found primarily in the referral hospital and university wastewater. The rare ARGs or those at individual sites included *blaCARB-4* and *aadA17* in university wastewater. The ARGs exclusively found in the hospital wastewater included tet*(G), tet(Q), tet(40), qnrs2, nimE, mph(G), floR, erm(T), erm(A), dfrG, dfrA1, catB3, blaOXA-97, blaNDM-1, blaGES-2, blaCMY-4, blaCARB-2, blaCARB-4, ARR-3, ARR-6* and *aadA1* (Figure 2). No rare ARG genes were present in the influent or effluent of the WWTP. We also identified several clinically relevant antibiotic genes in addition to the core antibiotics identified across the different wastewater sites. For example, hospital wastewater contains several relevant clinical beta- lactamase genes, such as *blaOXA* (*blaOXA-10, blaOXA-58, blaOXA-97*), *blaNDM-1, blaCMY-4, blaGES-2* and *blaCARB-2,* while university effluent contains *blaOXA-10* and *blaCARB-4* (Figure 2). The tetracycline resistance genes *tet(A), tet(M),* and *tet(W)* were also found predominantly in the university effluent and referral hospitals (Figure 2).

**Figure 2.**
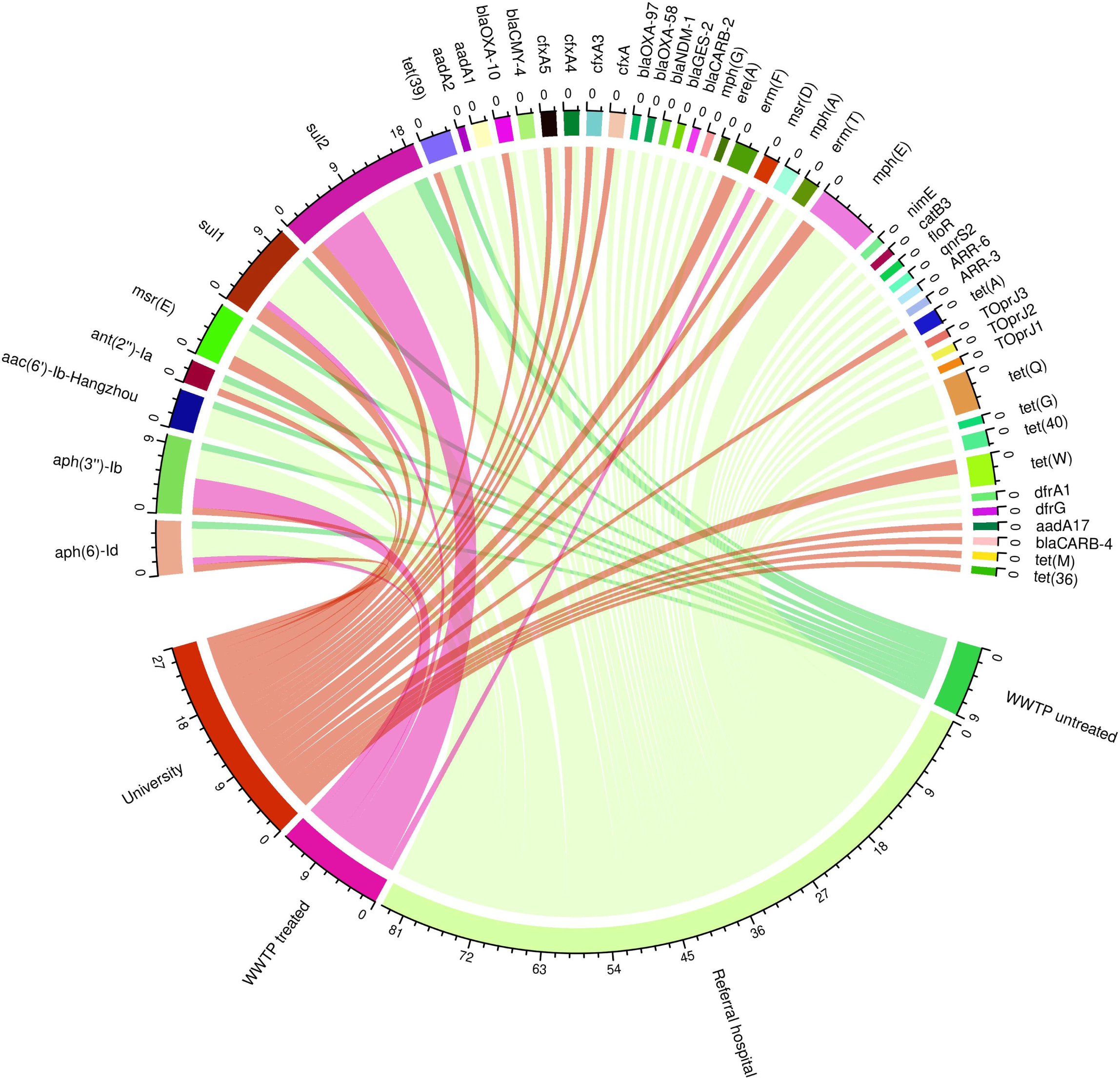
Circos plot showing the interactions of various wastewater ARGs across hospital, university and influent and effluent of a WWTP.

Analysis of the transcript abundances from wastewater collected from the referral showed high levels of *tet(G), msr(E),* and *mph(E)* and moderate expression of other ARGs associated primarily with tetracycline resistance (*tet(W), tet(M), tet (39), tet (36))*, and beta-lactams (*cfxA5, cfxA4, and cfxA3*). Low abundances of the beta-lactam *blaOXA genes* (*blaOXA-93, blaOXA-58, blaOXA-10*), *blaNDM-1, and blaGES-2,* as well as *Toprj1, Toprj2*, and *Toprj3, were also observed*. Wastewater from the university had higher transcript levels of *sul1* and *sul2* ARGs (Figure 3).

**Figure 3.**
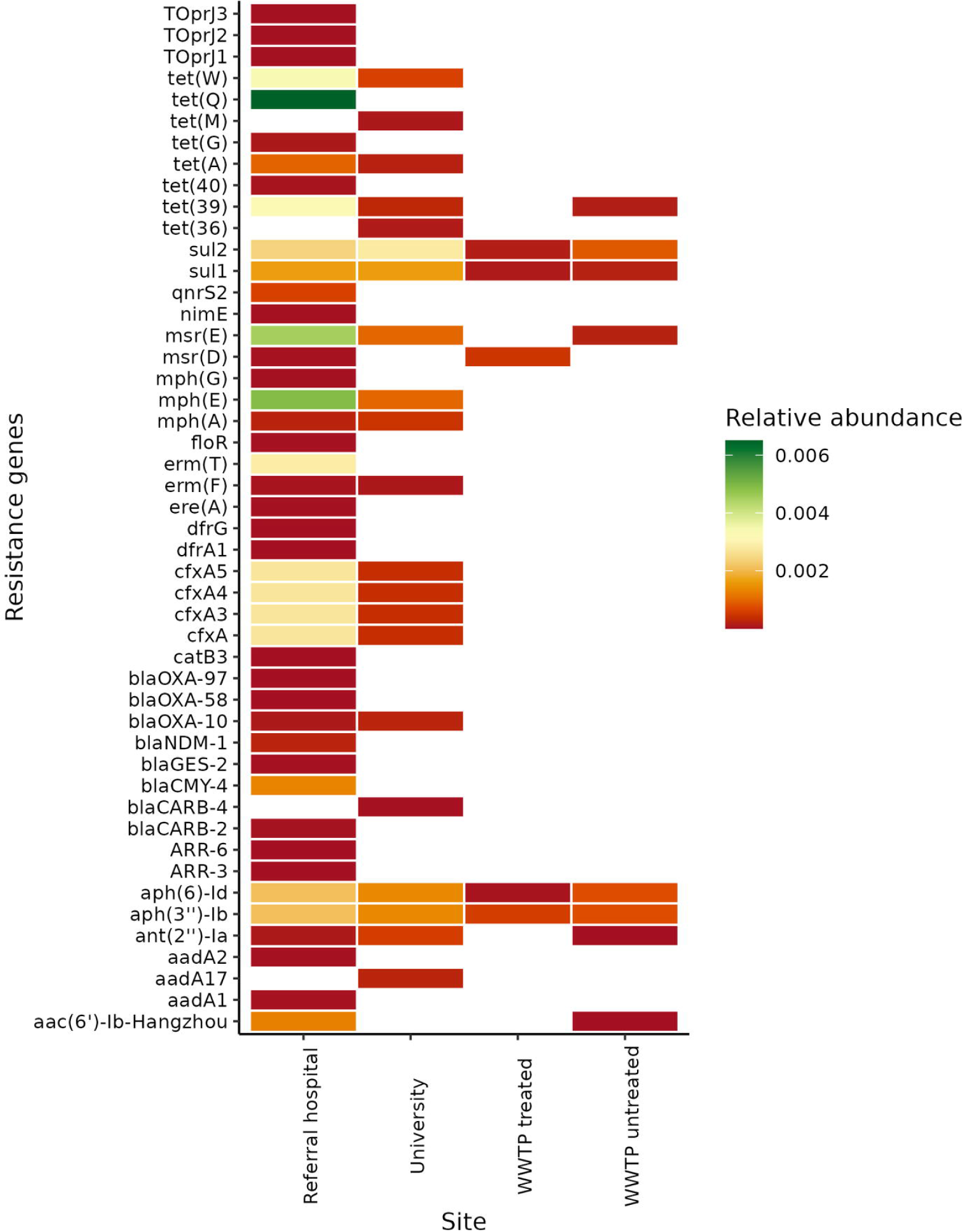
Total relative abundance of ARGs present in wastewater across different sites.

### 3.2 Identification of Mobile Genetic Elements and Virulence Factors

Plasmids and virulence factors were detected in wastewater from the hospital and university. The plasmids detected from the transcripts from the referral hospital included *Col(MG828), IncQ1, Col(pHAD28), IncW,* and *IncP1,* while those from the university wastewater included *ColRNAI, Col156, Col(pHAD28), IncFIB(AP001918)* and *Col(pHAD28).* In addition to plasmids, insertion sequences were also identified in hospital and university wastewater, with some being located in the same transcript as ARGs. The plasmid transcript *IncQ1* present in hospital wastewater was the same as the ARG *blaCMY-4* (Table 1). Insertion sequences within the same transcripts as ARGs were predominantly found in the hospital and wastewater samples (Table 1). Most ARGs in hospital wastewater were found in IS6, which was resistant to sulfonamides, aminoglycosides, and macrolides.

**Table 1.**
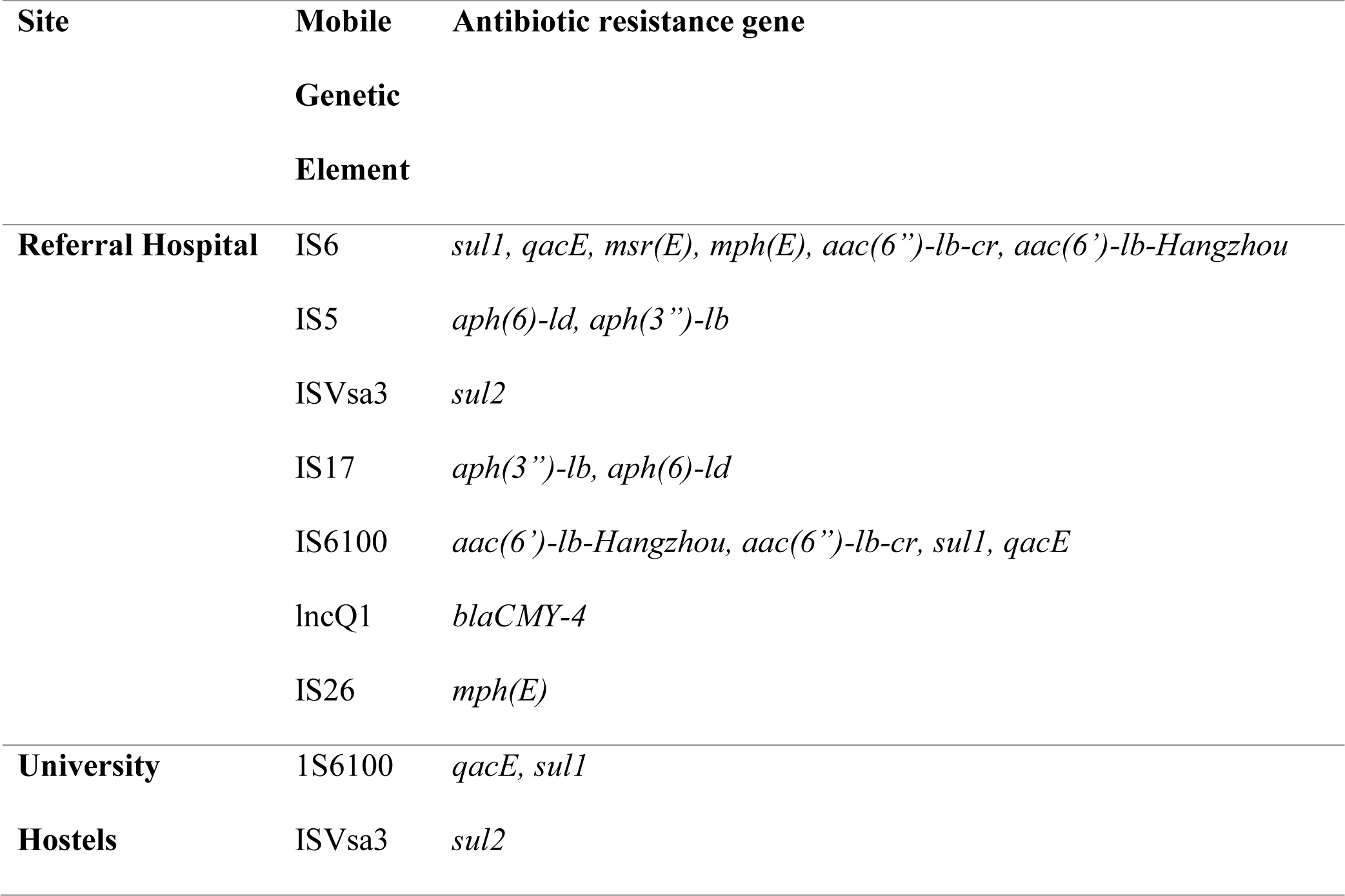
Antibiotic resistance genes found in the same transcript with mobile genetic elements.

Virulence factor transcripts detected from the hospital wastewater were associated with adherence (type IV pilus twitching motility protein *PilT*, twitching motility protein *PilT*, type IV pilus biogenesis protein *PilP*, and twitching motility response regulator *PilG*), motility (chemotaxis protein *CheY*, flagellar export protein *FliJ)*, immune modulation (outer membrane protein *OmpA*), and effector delivery systems (general secretion pathway protein G and general secretion pathway protein I). The virulence transcripts detected in the university wastewater were primarily associated with motility (flagellar basal body L-ring protein, flagellar biosynthesis protein FliO, flagellar motor switch protein FliN, and chemotaxis-specific methylesterase) and adherence (pili biogenesis protein MshG GspF- like).

### 3.3 Taxonomy classification

We also identified bacterial taxa across different wastewater sites. Across all four sites, *Gammaproteobacteria* was the predominant class of bacteria present. However, the hospital wastewater also contained *Bacteroidota and Bacillota,* while *Bacteroidota* was a major class in the WWTP influent (Figure 4A). At the genus level, hospital wastewater was primarily composed of *Acinetobacter, Aeromonas, Bacteroidales, Klebsiella,* and *Uruburuella,* while university wastewater contained *Aeromonas, Acinetobacter, Bacteriodales, Klebsiella,* and *Flavobacteriales. Aeromonas* and *Carnobacteriaceae* were the predominant genera detected in the WWTP influent, while *Acinetobacter* and *Aeromonas* composed a large proportion of the WWTP effluent (Figure 4B). We further sought to identify WHO-priority pathogens from the different wastewater treatment sites. *E. coli* made up the largest portion of WHO priority pathogens from wastewater present in the hospital, university and WWTP influent, while *Acinetobacter baumannii* had the highest proportion in the WWTP effluent (Figure 4C). Other WHO priority pathogens identified included *Helicobacter pylori, Neisseria gonorrheae*, *Enterococcus* spp., and *Pseudomonas aeruginosa (*Figure 4C).

**Figure 4.**
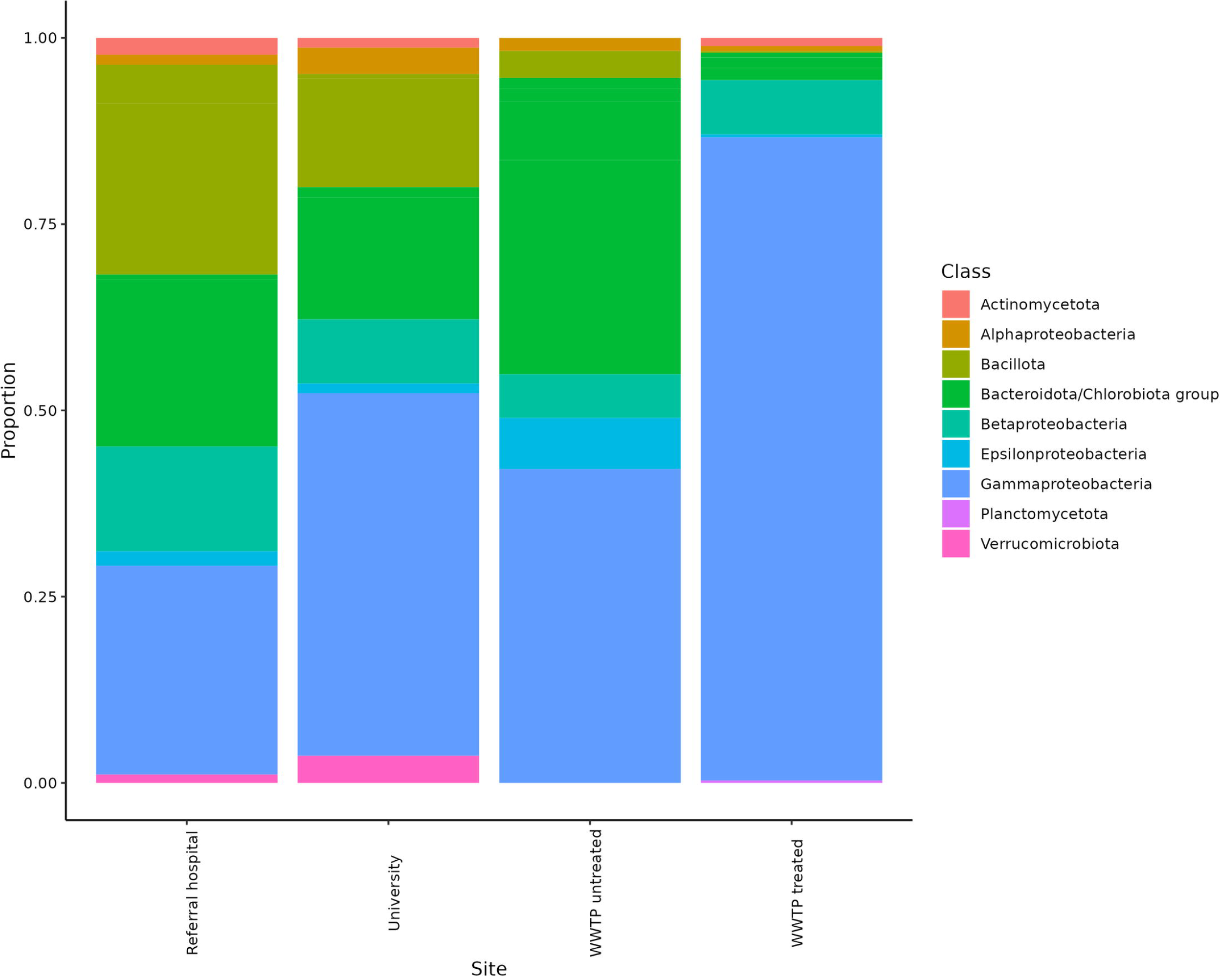
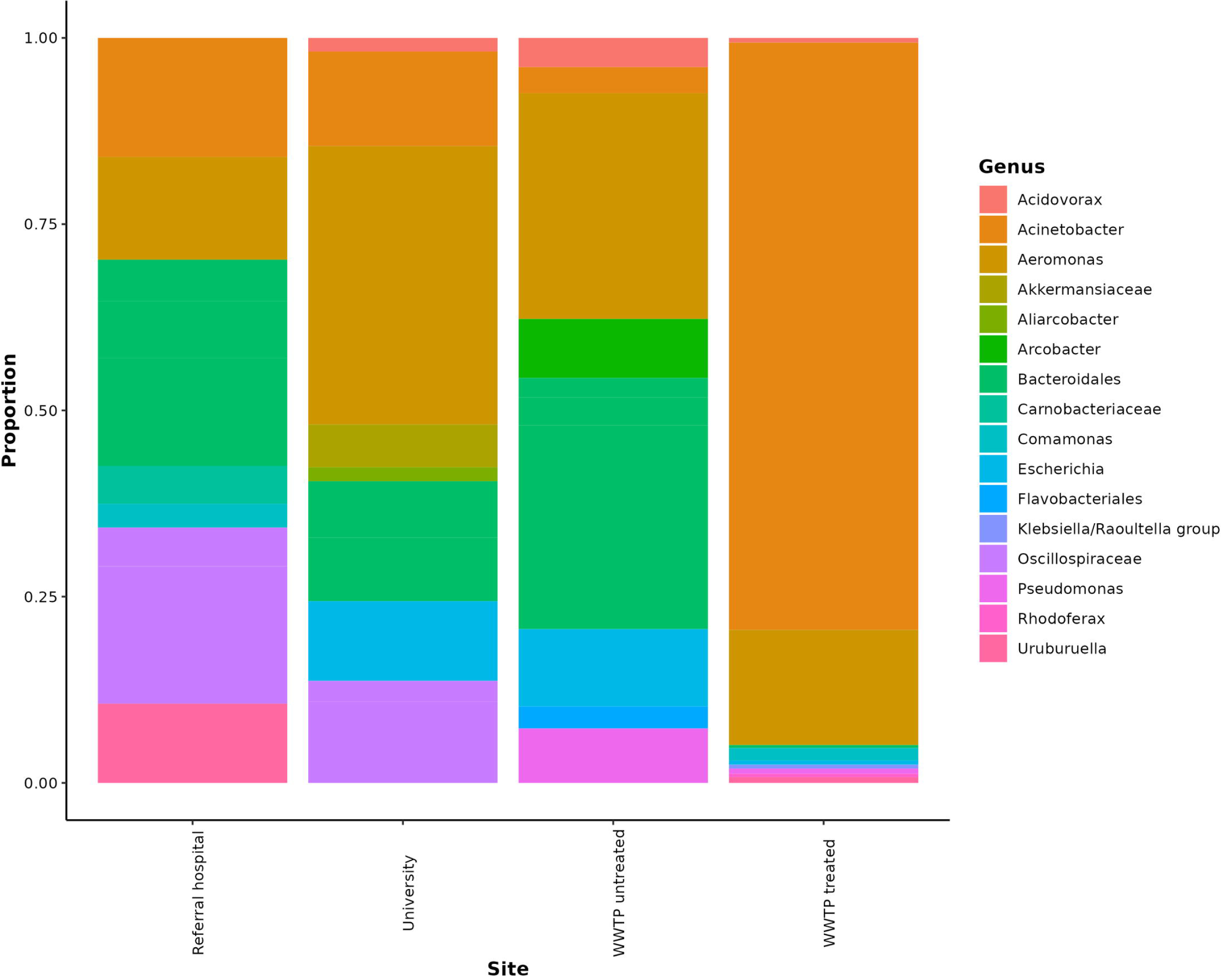
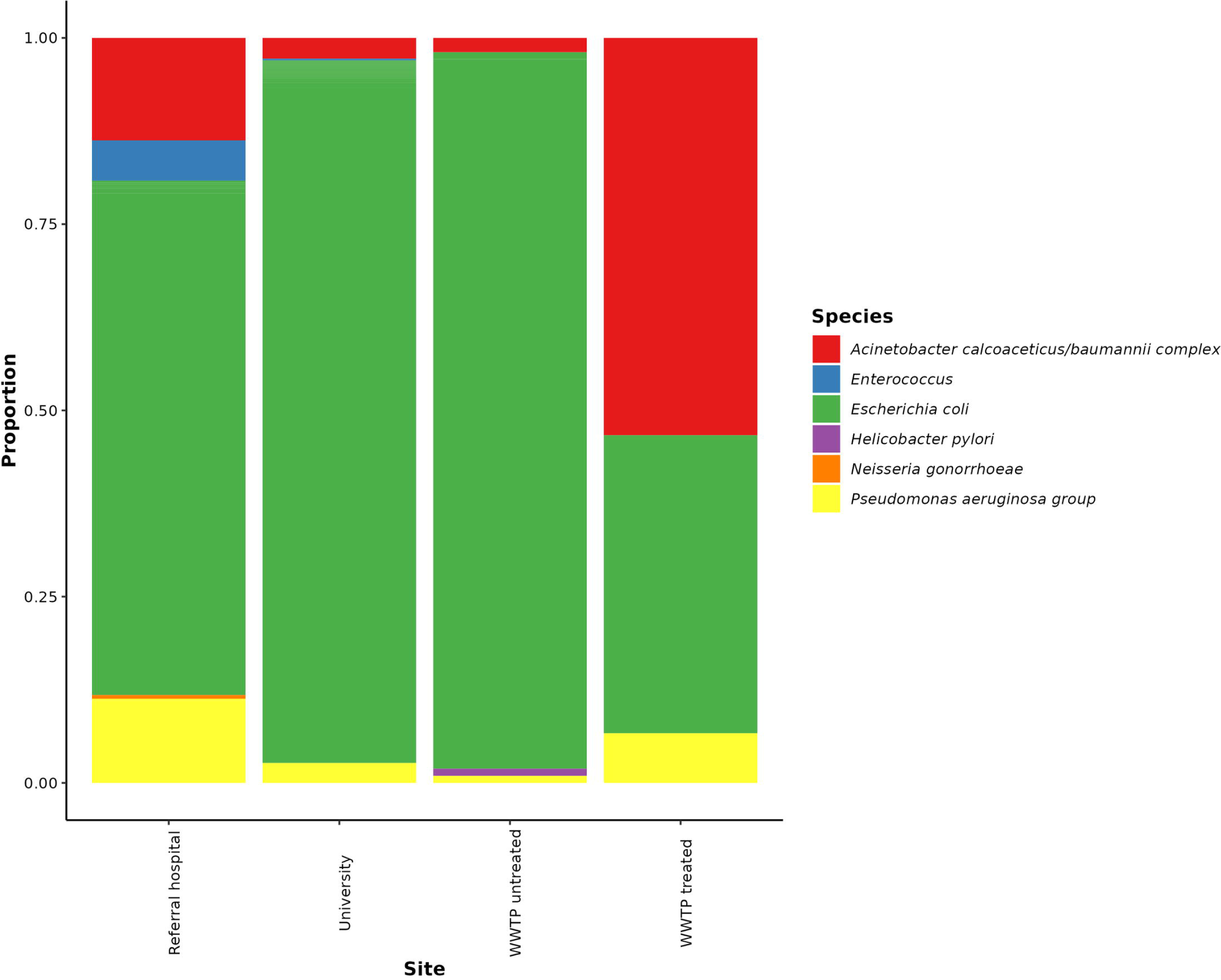
Taxonomic classification of bacterial organisms present across various wastewater sites. **A)** Proportion of bacterial organisms at the class level. **B)** Proportion of bacterial organisms at the genus level. **C)** Proportion of WHO list of priority pathogens identified from different wastewater sites after taxonomy classification using Kraken.

## 4. Discussion

Wastewater surveillance is a valuable method for monitoring the presence of ARB and the overall resistome. Our current study sought to identify ARGs from various wastewater samples. Tracking the spread of ARB and ARGs from very high-risk areas, such as wastewater from hospitals, universities and the influent and effluent of WWTPs, is critical for obtaining information used to track the spread of ARGs to the environment.

Our results revealed a greater number of ARGs in referral hospital wastewater than in the university, influent, or effluent of WWTPs. These large quantities of ARGs from hospital wastewater could be attributed to the frequent use of antibiotics within hospital settings and higher proportions of resistant bacteria and antimicrobial resistance genes [26,27]. Hospital wastewater contains a large abundance of ARGs and remains a hotspot for disseminating antimicrobial resistance genes to the environment by favoring the growth of bacteria harboring resistance genes via mobile genetic elements. These bacterial organisms survive and thrive in environments under selective pressure and can potentially transfer ARGs to other bacterial organisms that could be pathogenic to humans. Similarly, the high number of ARGs present in university settings could be linked to the misuse of antibiotics outside the hospital setting. The low number of ARGs in the influent and effluent of WWTPs could be attributed to the dilution of ARGs. Notably, the presence of various ARGs against sulfonamides, aminoglycosides, and macrolides suggested that these ARGs were not eliminated during the treatment process, as previously reported in other studies [8,10]. Our results confirm the need to continuously assess methods used to treat wastewater before it is ultimately released into the environment, considering that treated wastewater is released for use via various channels. Otherwise, there is a potential risk of introducing these ARGs into normal food chains through irrigation, further exacerbating the threat that AMR poses.

Our analysis identified a set of ubiquitous core ARGs across the different wastewater treatment sites. Sulfonamide ARGs have been detected in wastewater effluents at varying concentrations independent of the environmental conditions [28,29]. The identified core ARGs have also been previously reported when assessing the global resistosome [30]. The presence of these core ARGs, such as *sul1, sul2*, *aph(6)-ld,* and *aph(’3’)-lb,* could be linked to prolonged clinical use of sulfonamides and aminoglycosides both within and outside clinical settings. Multiple studies have also reported the most abundant ARG gene identified*, tet(G),* from hospital wastewater together with other tetracycline genes, attributing this difference to the overuse or misuse of antibiotics [31,32]. These core antibiotics could be used to monitor the spread of ARGs across different sites before they are released into the environment.

Several clinically relevant ARGs, including *blaNDM-1, blaOXA-10,* and *bla-OXA58,* were also detected in the hospital effluent. The presence of these clinically relevant bacteria in wastewater collected from hospitals may provide information about the increase in resistance to beta-lactam antibiotics. Moreover, these findings could lead to the use of an early warning system for the emergence or spread of multidrug-resistant strains, especially if similar ARGs are identified in clinical isolates. *blaNDM-1* shows resistance to multiple different classes of antibiotics, including aminoglycosides, beta-lactams, and fluoroquinolones, increasing its resistance to multiple agents [33]. The presence of *blaNDM-1* in hospital wastewater has also been shown in other studies [34,35].

The horizontal transfer of ARGs is a common dissemination method for antimicrobial resistance. Our analysis revealed the presence of multiple insertion sequences in the same transcripts as ARGs. Insertion sequences are mobile genetic elements present in bacterial genomes and contribute to AMR by relocating resistance genes to different locations in the genome and promoting their transfer between bacteria. IS6 family insertion sequences, including IS26 and IS6100, were identified from hospital wastewater. These insertion sequences are commonly found in gram-negative bacteria in association with ARGs and could constitute one of the methods by which ARGs are spread to the environment from wastewater [36]. These findings are consistent with previous reports on hospital wastewater, which also linked the potential role of IS6 family insertions in facilitating and expressing ARGs

[37,38]. Thus, the insertion sequences reported in this study could play a role in the horizontal gene transfer of ARGs against sulfonamide, aminoglycoside, and macrolide genes, while the identified plasmid could aid in the transfer of a beta-lactam ARG. In addition to mobile genetic elements, hospital and university wastewater contains virulence factors associated with adherence, immune modulation, and motility, which assist bacteria in competing for existing resources and surviving in their current environment.

Taxonomic classification allows the identification of various bacterial populations in wastewater sites. Some identified genera, such as *Aeromonas,* are part of the bacterial population predominantly found in wastewater [39]. Apart from the normal bacterial populations present in wastewater, we further identified WHO priority pathogens, including *N. gonorrheae*, *E. coli, P. aeruginosa*, *A. baumannii,* and *H. pylori,* from various wastewater sites. The presence of some of these WHO-priority bacterial strains could further spread ARGs, which are potentially harmful to human and animal life. Although short reads do not completely reveal which ARG transcripts are linked to specific bacterial genomes, recent advances in long-read sequencing are expected to fill this gap.

Meta-transcriptomic analysis offers a promising approach for assessing the local resistome, especially from various wastewater sites, and examining whether WWTP treatment fully eliminates ARGs before they are released into the environment. Our results identified four core antibiotic genes, *sul1, sul2, aph(6)-ld,* and *aph(’3’)-lb,* which could be used as targets for monitoring antibiotic resistance. Furthermore, we identified several clinically relevant beta-lactam resistance genes associated with multidrug resistance in hospital wastewater. Finally, we identified several ARGs in WWTP effluent, suggesting the need to improve the treatment process, especially in eliminating ARGs and other potential pathogenic bacteria. Our results highlight the potential role of using wastewater in monitoring the spread of ARGs to the environment, which could be adopted for AMR surveillance, especially within LMICs.

## Data Availability

All data produced are available online at https://www.ncbi.nlm.nih.gov/bioproject/?term=PRJNA1054104

https://www.ncbi.nlm.nih.gov/bioproject/?term=PRJNA1054104

## Acknowledgment

We acknowledge staff from Mount Kenya University, Thika Level 5 referral hospital and the Thika Wastewater Treatment Plant for their insights into this work.

